# Effect of different resumption strategies to flatten the potential COVID-19 outbreaks amid society reopens: a modeling study

**DOI:** 10.1101/2020.06.25.20140418

**Authors:** Yong Ge, Wenbin Zhang, Jianghao Wang, Mengxiao Liu, Zhoupeng Ren, Xining Zhang, Chenghu Zhou, Zhaoxing Tian

**Affiliations:** State Key Laboratory of Resources and Environmental Information Systems, Institute of Geographic Sciences & Natural Resources Research, Chinese Academy of Sciences, Beijing 100101, China; University of Chinese Academy of Sciences, Beijing, 100049, China; Emergency Department of Peking University International Hospital, Beijing, China

## Abstract

The effect of the COVID-19 outbreak has led policymakers around the world to attempt transmission control. However, lockdown and shutdown interventions have caused new social problems and designating policy resumption for infection control when reopening society remains a crucial issue. We investigated the effects of different resumption strategies on COVID-19 transmission using a modeling study setting. We employed a susceptible-exposed-infectious-removed model to simulate COVID-19 outbreaks under five reopening strategies based on China’s business resumption progress. The effect of each strategy was evaluated using the peak values of the epidemic curves vis-à-vis confirmed active cases and cumulative cases. We found that a hierarchy-based reopen strategy performed best when current epidemic prevention measures were maintained save for lockdown, reducing the peak number of active cases and cumulative cases by 50% and 44%, respectively. However, the modeled effect of each strategy decreased when the current intervention was lifted somewhat. Additional attention should be given to regions with significant numbers of migrants, as the potential risk of COVID-19 outbreaks amid society reopening is intrinsically high. Business resumption strategies have the potential to eliminate COVID-19 outbreaks amid society reopening without special control measures. The proposed resumption strategies focused mainly on decreasing the number of imported exposure cases, guaranteeing medical support for epidemic control, or decreasing active cases.

## Introduction

Cases of pneumonia with unknown origins were detected in Wuhan, China, in late December 2019 [1]. WHO announced the epidemic as an international public health emergency, naming it COVID-19 [2-3]. Although China has used a series of non-pharmaceutical strategies (including the Wuhan travel ban on January 23, 2020 [4-6]) to limit the epidemic, the high transmission ability of the coronavirus has led to secondary outbreaks worldwide [7-10]. Governments and the general public have expressed significant concern regarding these outbreaks and, at the end of May 2020, over six million confirmed cases and at last 300 thousand deaths have occurred due to this disease worldwide [11].

Contrasting the explicit life-threatening nature of COVID-19, non-pharmaceutical measures to curb the pandemic have also indirectly influenced people’s lives. China has kept its confirmed active COVID-19 cases at a mild order of magnitude, becoming the best example of pandemic control [12]. The rapid increase of confirmed cases in many other countries has driven their governments to set various strict pandemic prevention measures [13-16]. Such policies (e.g., lockdown and shutdown) have urged hundreds of millions of people to stay at home, creating new social problems (e.g., food shortage, increased unemployment, economic downturn) [17-21]. Given such circumstances, policymakers must cope with the tradeoff between strict epidemic preventions and social operation to address COVID-19 transmission and possible future outbreaks.

As COVID-19 infection has been eased with lockdown measures, some societies have suspended operations. China has reported a year-on-year fall in economic output for the first time since 1992, when the National Bureau of Statistics started releasing GDP growth numbers quarterly [22]. China’s service industry reported a 5.2 percent fall in value-added due to lockdown measures [23]. Despite this, the Chinese economy has recovered well with factories reopening and some workers returning to cities for work after the extended break caused by COVID-19 [24]. In addition, many other countries have also unveiled plans to reopen their societies, such as the US suggesting its states determine their own tactics according to local situations and proposing a phased strategy [25]. The UK has also unveiled a three-step plan to reopen its society [26]. However, these phased plans differ regarding the theme of each stage, as the US has prioritized industries (such as catering, entertainment, and religion) during its first phase while the UK has suggested those unable to work from home should return to the workplace during the first step. These differences are caused by various national or cultural circumstances, yet both plans emphasize that control measures (e.g., social distancing) should be maintained.

Investigating the impacts of the different business resumption strategies on COVID-19 transmission can provide critical information for reopening a society. In this study, we shall use observed epidemic data from Beijing to study five economic strategies in terms of China’s business resumption: direct reopening (DR), risk-based reopening (RR), order-based reopening (OR, theme-based reopening (TR) and a hierarchical reopening (HR). To model outbreaks under different strategies, the susceptible-exposed-infectious-removed (SEIR) model was employed in the simulation. As each resumption strategy considered no further control measures, we divided infectious subjects into undetected infectious (Iu) and detected infectious (Id) subjects, where the detection rate captured the current measures. The model parameters in our study have been calibrated by the observed epidemic data from Beijing with a reproductive number of 2.2, which is the evaluation of reproductive number for the initial COVID-19 outbreak in Wuhan. For stronger context, three reproductive numbers (1.4, 2.2, and 3.9) were compared, to represent stricter measures (1.4), and the partial lifting of current measures (3.9). The effects of each strategy were evaluated using the peak values of their epidemic curves vis-à-vis confirmed active cases and cumulative cases. Besides, spatiotemporal heterogeneity exists in the distribution of COVID-19 in worldwide [27-28]. Several studies confirm that the spread of COVID-19 is location dependent [29-30]. At different stages of resuming work, population flow is different driving spatiotemporal distribution of COVID-19 [31-33]. Thus, each district of Beijing is studied based on the local conditions.

The analysis of the work-resumption patterns documented in this article focused primarily on China as provides accurate information on the impacts of socioeconomic activities being resumed after COVID-19 transmission. In fact, China’s financial data saw a considerable rebound in March 2020, with business resumption nearly complete while the pandemic remains under control [34-36]. This study seeks provide insight into the impacts of business resumption on COVID-19 transmission, producing valuable information for other regions awaiting economic recovery in a post-pandemic world.

## Materials and methods

### Resumption strategies

Despite lackluster GDP numbers, economic activities have been recovering in China since early March, as virus transmission has been mostly under control since then [24]. Resumption progress has been seen in monthly indicators that include industrial production, retail sales, and fixed-asset investment. Reviewing China’s progress (Fig 1), allows resumption strategies to be summarized in five categories for exploring the epidemic transmission of COVID-19 in Beijing.

**Fig 1:**
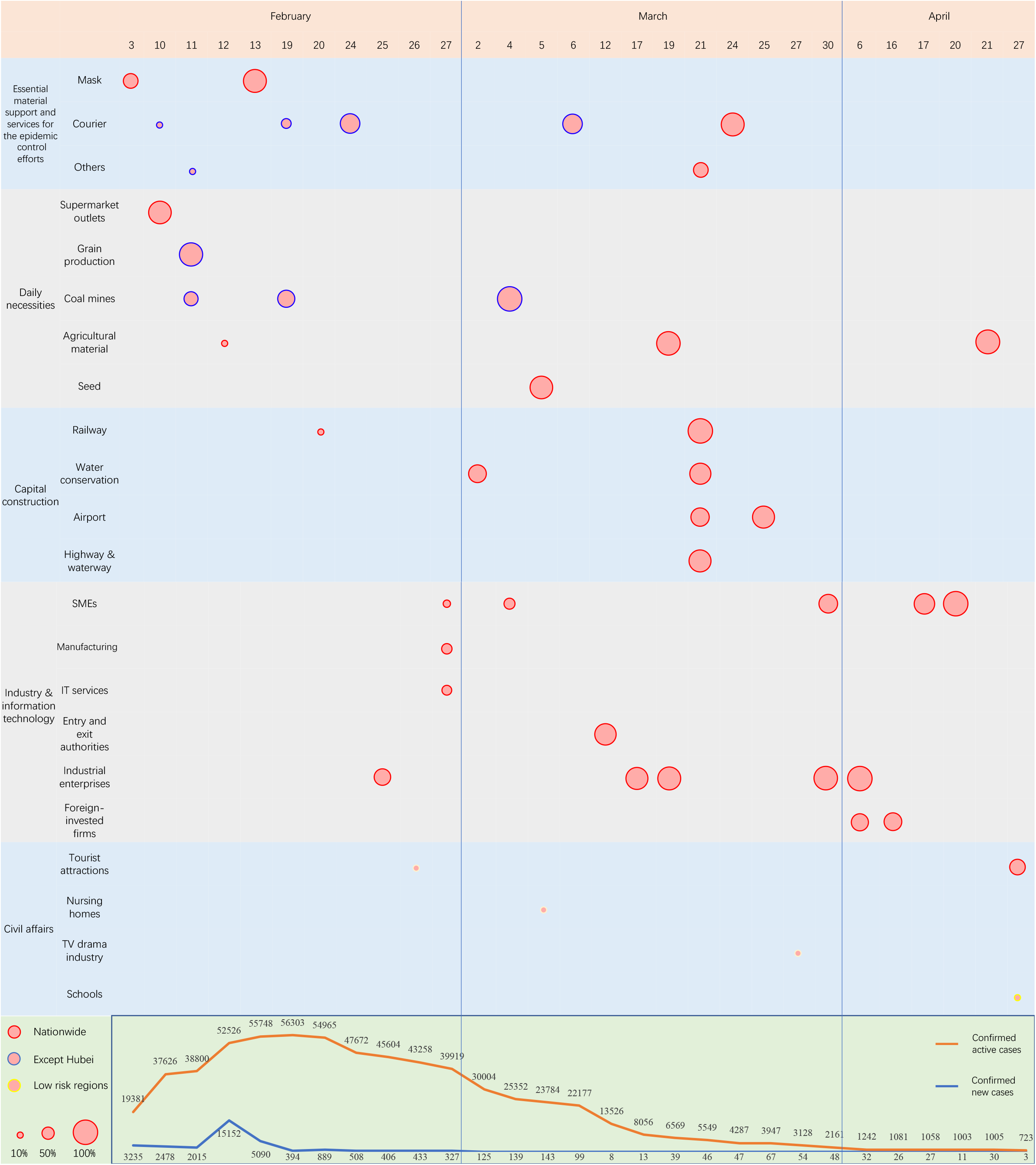
The progress of China’s business resumption.

China reopened its economy chronologically using five themes: Essential material support and services for epidemic control efforts, daily necessities, capital construction, industry, and civil affairs. The resumption progress of the first theme was reported on February 3, 2020, with China’s domestic mask producers resuming 60 percent of production capacity as the epidemic outbreak was developing. The other themes reported subsequently according to outbreak circumstances. For instance, civil affairs resumed when the epidemic had been somewhat curbed. The resumption ratio is characterized by circle size, and the resumption extent is depicted by circle envelopes (Source: see appendix).

### DR

The most straightforward strategy where the whole working population returns to the workplace together, and epidemic transmission is associated with no further disturbances.

### RR

In this strategy, only people living in low-and-moderate-risk regions are allowed to return to work. Such regions are defined as somewhere the COVID-19 incidence possesses a controlled level. In contrast to DR, the ratio of exposed cases to migrants decreased.

### OR

A more cautious approach would be to have people return to work in batches instead of all at once during the initial stage. We simplified this strategy by allowing one quarter of total migrants to return to work with a second quarter returning after three weeks. We did not distinguish the occupations of those resuming work.

### TR

Considering epidemic intervention, essential material support and services for epidemic control usually reopen first. After that, migrants who returned to work are assumed to have a reduced COVID-19 incidence because of sufficient medical material and services support. After three weeks, daily necessities reopen followed by infrastructure construction, industry, and civil affairs at last.

### HR

Comprehensively, TR allows migrants return to work should be of low- and moderate-risk regions, just as RR does. In this way, all migrants possess a lower COVID-19 incidence, which contrasts with TR, regardless of occupation.

For each theme, we separated the corresponding population equally into three groups with each group returning to work on subsequent weeks (OR).

### Modeling analysis

We captured the epidemic spread amidst work resumption based on SEIR dynamics [37], categorizing infections as Iu and Id subjects. Each infection detected is assumed to be wholly hospitalized, setting Id at zero for producing further transmission:

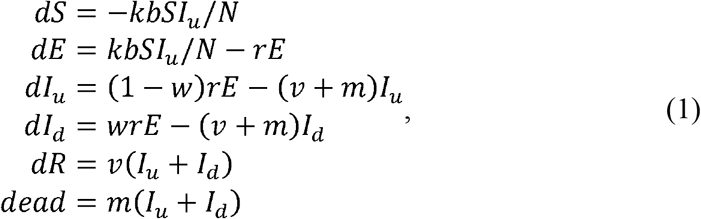

where *R*_0_ is the basic reproductive number, and *N* represents the total population. The basic reproductive number *R*_0_ *= kb*/(*w* + *v* + *m*) when there is no intervention, allowing *k* to represent the daily contact number per capita, and *b* to represent the probability of getting infected by per contact [38]. In our model, we replaced *kb* with *R*_0_(*w*+ *v* + *m*), because the intervention is characterized by *w*. As epidemic prevention measures are always positive to epidemic control, the model is not shaped by any control measures. Those returning to work are deemed asymptomatic due to related measures and are isolated from residents, so the only imported exposed cases come from returned workers, and the susceptible population remained limited to health workers. Therefore, epidemic evolution depended on resuming patterns of socioeconomic activities with a particular transmission model.

We ran 500 simulations for each strategy, and imported the exposed cases from a Poisson distribution with the expectation that exposed cases corresponded with the defined ratio for each stage. The model was calibrated using epidemic data from Beijing (cumulative cases, recoveries, and deaths) with a fixed *R*_0_ = 2.2, where the daily detection rate *w* reflected the epidemic prevention measures. For generalization, we assumed the imported exposed cases were proportional to the population of returned workers with a basic ratio of 0.1%, wherein the population of migrant residents evaluated the population of returned workers. The ratio for low- and moderate-risk regions was set to 70 percent of the basic ratio, and the restarting of essential material support and services for the epidemic control effect could further reduce COVID-19 incidence by 20%. The parameters of the model are given in Table 1.

**Table 1.** Parameters for the model.

|  | Explanation | Value | Reference |
| --- | --- | --- | --- |
| $R_0$ | The basic reproductive number | 2.2<br>(95% CI, 1.4 to 3.9) | Li et al. [39] |
| $1/r$ | The mean incubation period | 5.2 | Lauer et al. [40]<br>Backer et al. [41] |
| $w$ | The daily infection detection rate | 0.5762<br>(95% CI, 0.2655 to 0.8870) | Calibrated by epidemic data in Beijing |
| $v$ | The daily infection recovery rate | 0.0417<br>(95% CI, 0.0363 to 0.0471) | Calibrated by epidemic data in Beijing |
| $m$ | The daily death rate | 0.0009 | Wang et al. [42] |

## Results

Overall, as the reproductive number increased, the risk of the virus spreading increased under all five proposed business resumption strategies. DR represented natural outbreaks leading to most severe outbreak with the corresponding transmission being set as the baseline. Among the other four resumption strategies, HR was the most effective way to reduce COVID-19 incidence for reproductive numbers 1.4 and 2.2 (see Fig 2). Given a sizeable reproductive number of 3.9, the business resumption strategy had little effect.

**Fig 2.**
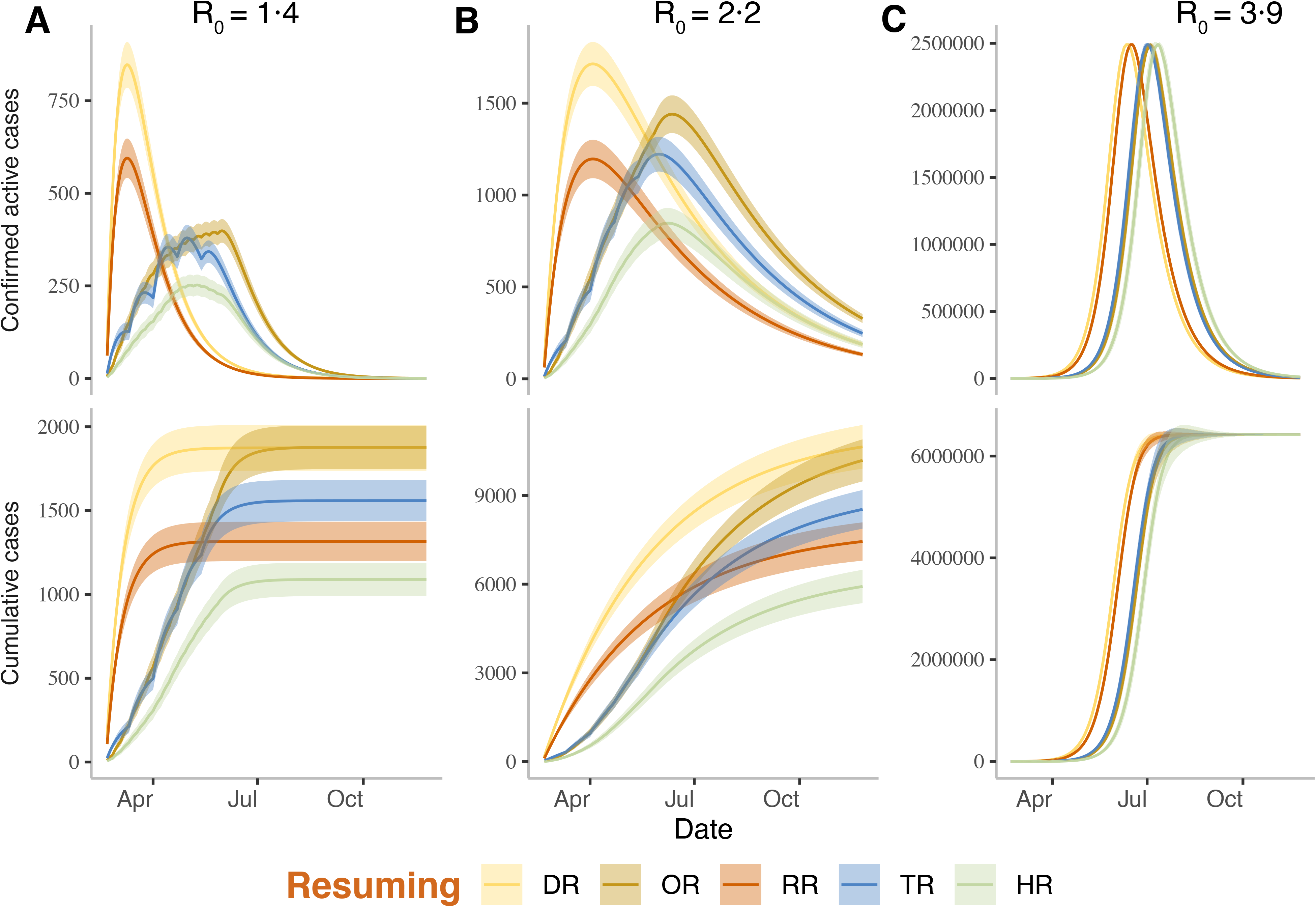
The simulated outbreaks amid different strategies of business resumption. (A) Cumulative and confirmed active cases for all five scenarios with a reproduction number (R0) of 1.4. (B) Cumulative and confirmed active cases for all scenarios with a reproduction number of 2.2. (C) Cumulative and confirmed active cases for all scenarios with a reproduction number of 3.9.

Confirmed active cases described how many infections were hospitalized, indicating strain on local medical services. The whole four strategies (RR, TR, OR and HR) performed well given a low reproductive number, while their effects decreased as reproductive numbers increased. Cumulative cases represented infected subjects including imported exposed cases. Given consistent epidemic prevention measures, OR could not decrease the cumulative cases (Fig 2). Our simulation set February 20, 2020, as the day for reopening society. With a reproductive number of 1.4, DR and RR controlled transmission within one month (by early April, 2020), while OR, TR and HR required four months (up to late June) to control the epidemic. Given a reproductive number of 2.2, each strategy required roughly nine months (up to late November of 2020) to control the epidemic. A reproductive number of 3.9 saw each strategy needing about five months (until early July, 2020) to control the epidemic. While lifting the control measures (*R*_0_ = 3.9) could lead to a shorter COVID-19 life than maintaining current control measures (*R*_0_ = 2.2), the height of the peak for confirmed active cases (hospitalized) under each strategy was considerably higher than that of *R*_0_ = 2.2, indicating the COVID-19 curve was not flattened.

Four business resumption strategies led to reduced COVID-19 incidence with differing effects, as shown in Fig 3. In the simulation, RR eased the epidemic by decreasing the total imported exposed cases according to regional COVID-19 incidence (i.e., 70% of the DR strategy). OR eased the epidemic by reducing daily infections and, consequently, the total number of imported exposed cases remained identical to DR. TR functioned equivalently to OR as it also asked people to return to work in batches by occupation. As this strategy called epidemic prevention occupations to return first, it was assumed that regional COVID-19 incidence would decrease accordingly. Therefore, TR imported 75% of exposed cases compared to DR and, to this end, HR combined the above three strategies to grant it the lowest number of imported exposed cases (52.5% of DR).

**Fig 3.**
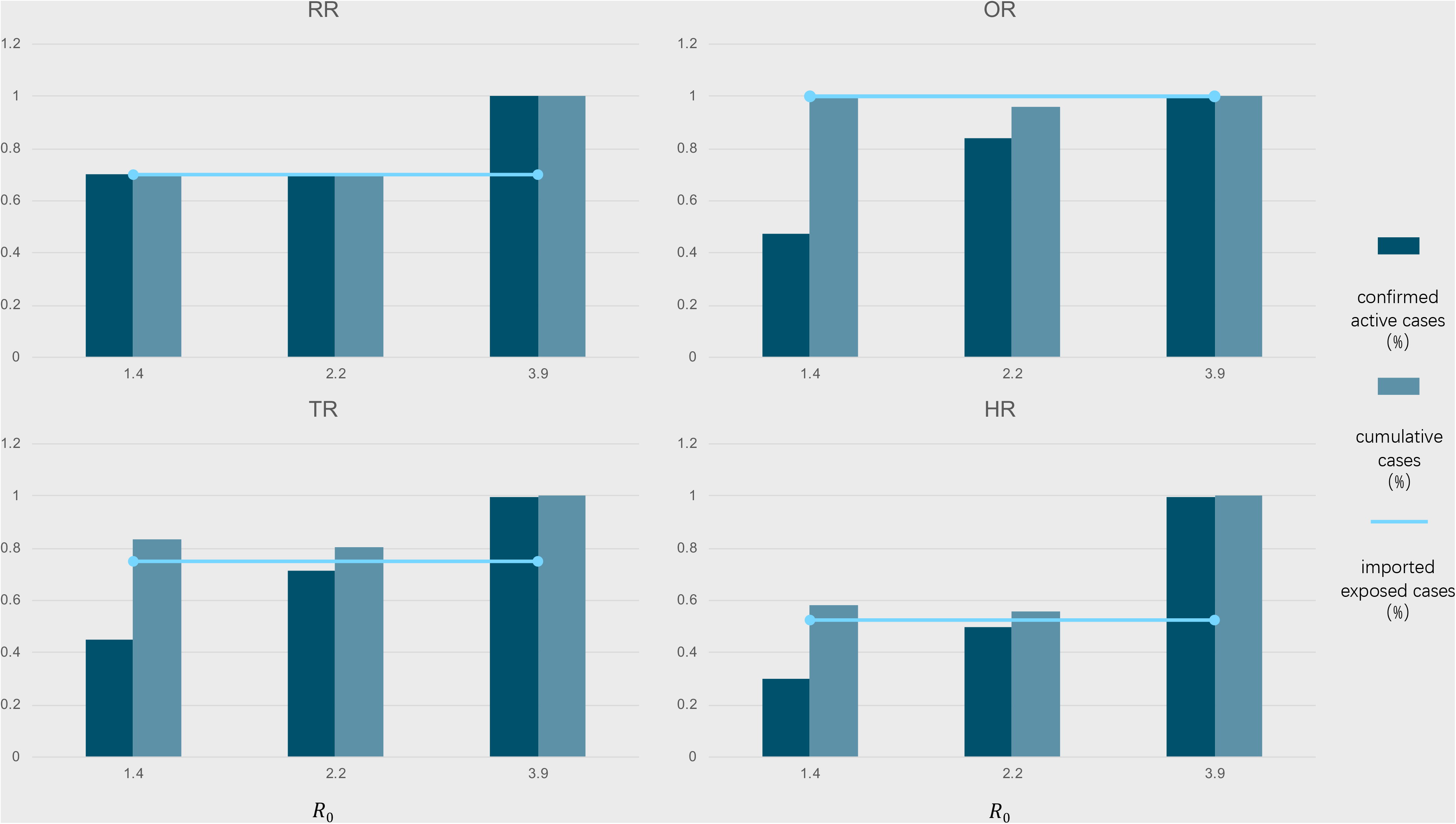
Strategy effects for COVID-19 prevalence reduction measured against direct resumption.

With stricter control measures (*R*_0_ = 1.4), RR reduced peak numbers of confirmed active and cumulative cases to about 70% of DR. OR only reduced cumulative cases to 47% of DR. TR reduced peak numbers of confirmed active and cumulative cases to about 44.9% and 83.2% of DR, respectively. HR again achieved the greatest reductions of 29.9% confirmed active cases and 58.3% cumulative cases to DR. When the reproductive number rose to 2.2, all strategy effects decreased slightly. HR still performed the most effectively, reducing peak numbers of confirmed active and cumulative cases to 49.64% and 55.88% of DR, respectively. As the sizeable reproductive number reached 3.9, all strategies only had slight COVID-19 prevalence reduction effects with all peak numbers being comparable to DR.

For each reproductive number, the heights presented the ratio of peak numbers between each strategy and DR, with respect to confirmed active, cumulative and imported exposed cases. The RR, OR, TR, and HR had 70%, 100%, 75% and 52.5%, respectively, of imported exposed cases compared to DR. Given a reproductive number of 1.4, peak numbers of confirmed active and cumulative cases for RR were about 70% of DR, while OR, TR, and HR reached (47%, 100%), (44.9%, 83.2%) and (29.9%, 58.3%), respectively. With a reproductive number of 2.2, HR performed best with (49.64%, 55.88%). With a reproductive number of 3.9, all strategies had little effect as all ratios were nearly 100%, showing a failure to reduce outbreaks with smaller numbers of imported exposed cases.

In addition to the resumption strategy, different type of workplaces associated with different risk of secondary outbreaks of COVID-19 amid society reopens. In general, the crowd-intensive place and the labor-intensive-place possess a higher risk. The crowd-intensive workplace refers to somewhere population flow is large, such as shopping mall, catering, and office buildings. While the labor-intensive workplace indicates someplace the labor force is, to some extent, isolated from the external environment. The situation of Beijing is illustrated in Fig 4, the crowd-intensive places are centralized in Xicheng Districts (XC) and Dongcheng District (DC), and radioactively diffuse to the periphery; and the labor-intensive places are positioned in the southern Beijing, especially in Shijingshan District (SJS), Fengtai District (FT), and Chaoyang District (CY). Much attention should be paid to the above districts due to their intrinsic risk of the secondary outbreaks of COVID-19 to reopen society.

**Fig 4.**
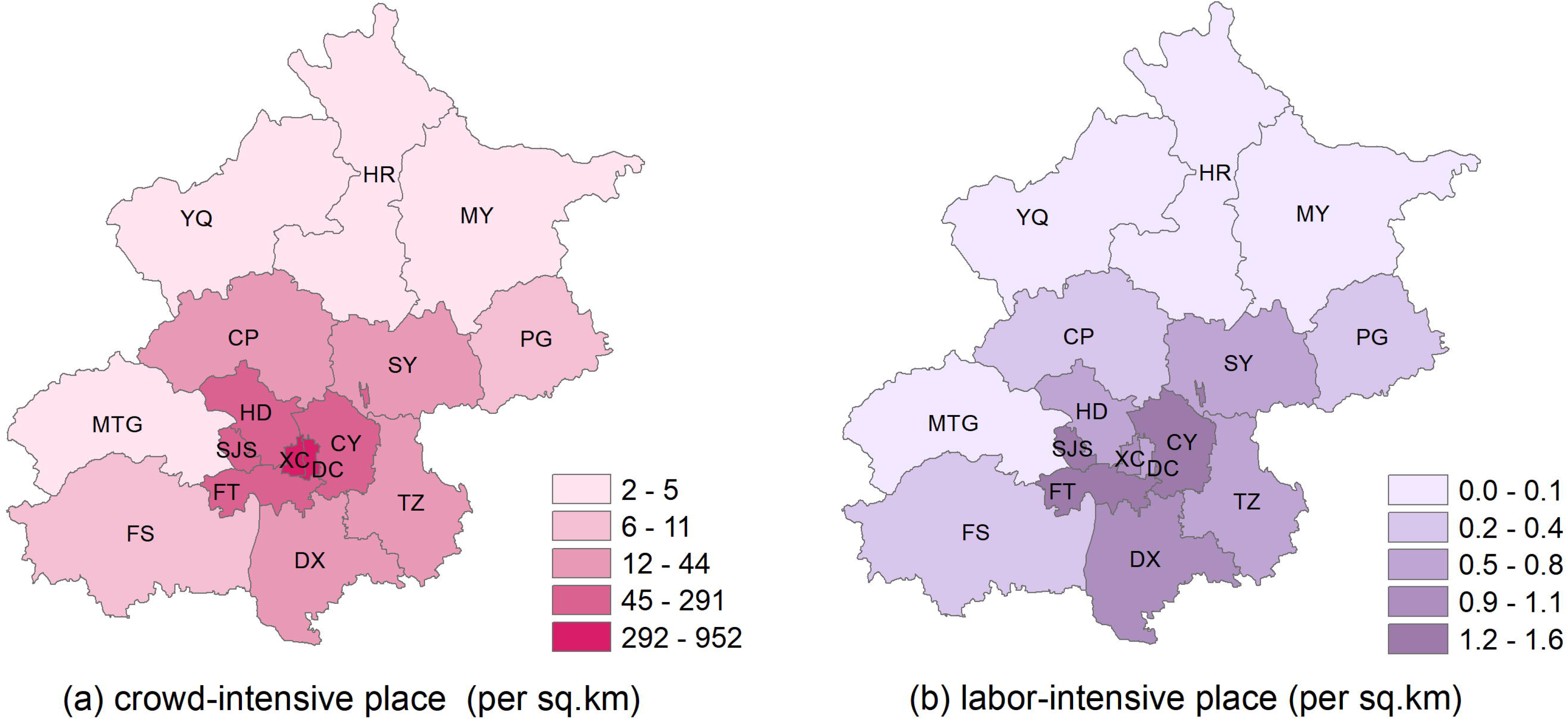
Spatial distribution of the crowd-intensive and the labor-intensive workplaces.

Moreover, the effects of strategies in this study were not sensitive to population scaling. Districts with larger populations are intrinsically high-risk regions as an economy reopens. It is reasonable to study the impact of local migrant resident populations on different strategy effects [43]. Therefore, we generated COVID-19 transmission results for three districts of Beijing, with small (48,000), moderate (306,000), and large (1,579,000) migrant residents with a reproductive number of 2.2 (Fig 5) [44]. Under three graduated population scales, corresponding to Yanqing District (YQ), Fangshan District (FS) and Chaoyang District (CY) respectively, the cumulative cases and confirmed active cases affected by hierarchy-based resumption (HR) with a reproductive number of 2.2 are shown on the left of the Figure 5. The effect of HR strategy is comparable for all three districts.

**Fig 5.**
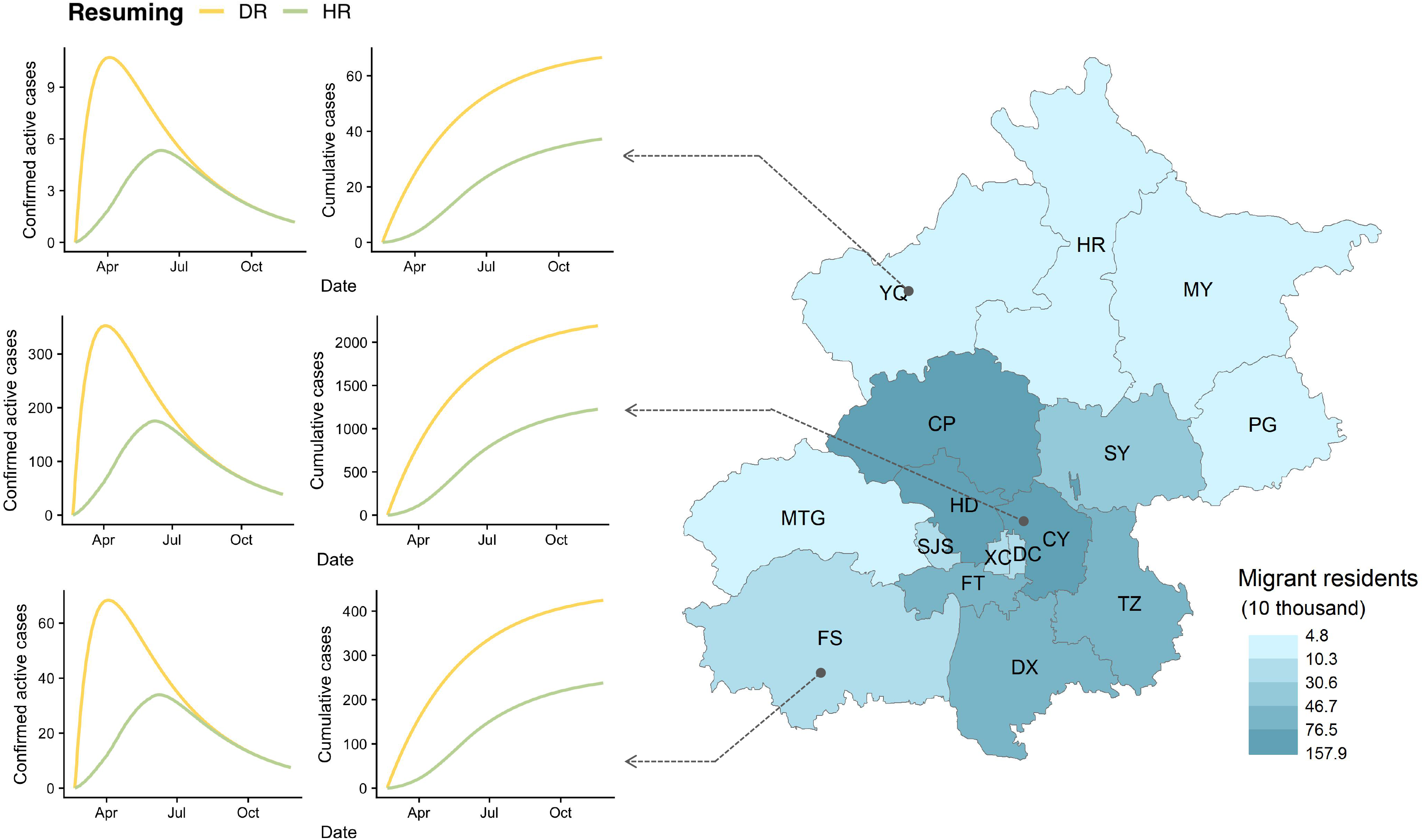
Impact of population-scale on the effects of hierarchy-based resumption.

## Discussion

We examined the effect of different resumption strategies to reduce COVID-19 prevalence amid societal reopening, capturing corresponding strategic influences on epidemic transmission. We found that OR, RR, TR and HR strategies could weaken an epidemic in terms of confirmed active cases with current epidemic prevention measures in Beijing. Using stricter measures than DR strategy found HR having the strongest effects. The lowest reproductive number in this study, 1.4, represented stricter epidemic prevention measures than those used for model calibration, while the highest reproductive number, 3.9, represented less intervention. Strategic effects decreased as epidemic prevention measures were lifted.

Our model did not include additional epidemic prevention measures that might have decreased COVID-19 transmission capability and, therefore, increase strategic effectiveness. The baseline reproductive number of our model was the first estimated reproductive number provided by Wuhan’s epidemic data. We also modified the standard SEIR model by dividing those categorized as infectious further into Iu and Id categories, with Id representing infections that were detected and subsequently hospitalized to isolate them and eliminate their transmission ability. Other parameters were calibrated using Beijing epidemic data, placing local control measures within these parameters. Adjusting the reproductive number employed in the simulation represented differing intervention states.

As one of the biggest cities in China, Beijing has 21.5 million residents, 7.6 million of which are domestic migrants [44]. Such hosts of imported workers make Beijing a high-risk region in terms of resuming work. For Beijing districts though, significant heterogeneity exists in population distribution and local socioeconomic factors. Our results show that districts with a high migrant density, such as the Chaoyang District, would witness more confirmed active cases and face higher risks of secondary outbreak. However, such districts may not become high-risk regions as the risk of resuming work is also based on each district population’s spatial heterogeneity characteristics, socioeconomic factors, prevention measures, and medical capacity. For example, most residents would stay home for much of the time if a district has few open work units.

Although China has almost completed its business resumption, there has not yet been a secondary COVID-19 outbreak. To characterize each strategic effect, we simply assumed one exposed case per ten thousand returned people. Only exposed cases were imported due to strict travel restrictions and control measures in place to prevent those with symptoms going to work. In this way, projected infections may not be consistent with China’s situation, but our simulations have shown that China’s business resumption model can still reduce COVID-19 prevalence with current control measures. Given small to medium reproduction numbers, a business resumption strategy has significant influence on COVID-19 prevalence reduction, particularly HR of socioeconomic activities. When reproduction increased to a sizeable number, business resumption strategy had a negligible effect. Besides, stringent measures should be taken by local governments in different regions based on the local conditions of each district. Regions with crowd-intensive places, such as shopping mall, catering, and office buildings, should pay special attention to strengthening air circulation and cleaning and disinfection. In labor-intensive workplace, daily temperature monitoring is encouraged as well as strict control the problem of excessive numbers of workers in confined rooms.

Our study indicates that societal reopening strategies can support epidemic control without further special control measures. As returning to work is an urgent need for several countries, this study could provide valuable insights into business resumption plans. Our model could also be calibrated according to national situations to test proposed plans based on local situation. However, migrants of different occupations are commonly placed in isolated workplaces, while our simulation simply placed them together. Transmission abilities may also vary with occupation, changing model parameters beyond our study’s limits.

## Data Availability

This study is mainly a simulation study. All data are available on the website of Beijing's CDC.

## Notes

### Competing Interest Statement

The authors have declared no competing interest.

### Funding Statement

There is no funding.

### Author Declarations

It is unnecessary for this study.

